# Comparison of frailty instruments in acute coronary syndrome patients

**DOI:** 10.1101/2023.11.06.23298187

**Authors:** Anne Langsted, Jocelyn Benatar, Andrew Kerr, Katherine Bloomfield, Gerry Devlin, Alex Sasse, David Smythe, Andrew To, Gerry Wilkins, Harvey White, Ralph Stewart

## Abstract

**Background:** Recognising frailty is important to guide clinical decisions in older patients with cardiac disease. The relative strengths of different frailty instruments to predict mortality and hospitalization risk are uncertain.

**Methods:** The Edmonton Frail Scale (EFS), Fried Criteria (Fried), Clinical Frailty Scale (CFS), Katz score (Katz), GP Cognition test (GPCog), and Euroscore II a disease based risk score, were completed in 1174 clinically stable inpatients >70 years of age admitted with an acute coronary syndrome to 5 New Zealand hospitals. Associations with all cause mortality (n=353, 29%) during a median follow-up of 5.1 (IQR: 4.6-5.5) years and hospitalization for > 10 days in the next year (n=267, 22%) were evaluated.

**Results:** There were graded associations between increasing frailty assessed by each tool and all cause mortality. For the EFS, which scores up to 17 points on different dimensions of frailty, hazard ratios for high (score 9-17, n=197) compared to low frailty (score 0-2, n=331) were 5.0 (95%CI: 3.4-7.4) for mortality, and 5.3 (3.4-8.3) for hospitalization. Discrimination for all-cause mortality according to Harrell’s C-index for each instrument were EFS 0.663, Euroscore II 0.654, Fried 0.648, CFS 0.640, GPCog 0.608, and Katz 0.593, P<0.001 for all. C-statistics for hospitalization >10 days were EFS 0.649, Fried 0.628, Katz 0.602, Euroscore II 0.589, CFS 0.584, and GPCog 0.552, P<0.001 for all. When combining tools integrated discrimination improvement for both mortality and hospitalization were greater for EFS than for other frailty instruments.

**Conclusion:** In acute coronary syndrome patients aged >70 years greater ‘frailty’ assessed using all tools was associated with higher mortality and hospitalization. The Edmonton Frail Scale, which provides a graded measure of severity of frailty based on information relevant to clinical care, discriminated the risk of mortality and hospitalization as well or better than other frailty instruments.

## Background

Frailty is characterized by multisystem impairment that decreases physiologic reserve and increases the vulnerability to stress(1). Frailty is known to be associated with lower quality of life, prolonged hospitalisations, and increased all cause mortality (2-4). The benefits of some cardiovascular interventions may be less in patients who are very frail (5). Clinical practice guidelines on management of acute coronary syndromes, heart valve disease, heart rhythm problems, and other cardiovascular diseases recommend frailty is considered when making treatment decisions for older patients(6-9). A formal evaluation of frailty may therefore be useful to identify patients at risk, and to help inform treatment decisions.

A number of different frailty assessment instruments are currently used. These include the Clinical Frail Scale (CFS) (10), the Fried criteria (Fried) (11), the Edmonton Frail Scale (EFS) (12) and Katz score (Katz) (13). Although cognitive impairment and dementia are distinct from frailty, tests for cognitive impairment such as the GP Cognition test (GPCog) (14) are also relevant to care of elderly patients with cardiovascular disease. The CFS is the simplest score – ranking overall frailty in 7 levels based on clinical judgement.

Differences between ‘frailty’ tools may be important to how frailty is interpreted clinically. Many studies have demonstrated associations between frailty assessed using different instruments and adverse clinical outcomes(2-4,15-17). However, few studies have directly compared how well different frailty instruments predict adverse clinical outcomes in patients with cardiovascular disease, or how they compare to standard disease based risk scores, such as Euroscore II (18).

The aim of this study was to compare how well several standard frailty instruments predict the risk of all cause mortality and prolonged hospitalisation during follow-up in older patients admitted to hospital with an acute coronary syndrome.

## Methods and materials

### Study Population

Patients aged more than 70 years admitted to a participating acute cardiac unit in New Zealand with an acute coronary syndrome were eligible for inclusion. Patients of Māori and Pacific descent could be included if >60 years, because age-related risk factors are present at a younger age in these peoples. 1231 patients were included from 5 New Zealand hospitals between August 4, 2015 and August 29, 2017. Frailty assessments were incomplete in 57 patients, so the study population is 1174 patients. This included 138 Maori or Pacific patients aged 60 to 70 years.

### Frailty asessments

Instruments assessed in this study were Clinical Frail Scale (CFS) (10), the Fried criteria (Fried) (11), the Edmonton Frail Scale (EFS) (12), the Katz score (Katz) (13), the GP Cognition test (GPCog) (14), and the Euroscore II (18). Types of questions included in each of the frailty instruments are summarized in Table 1.

**Table 1.**
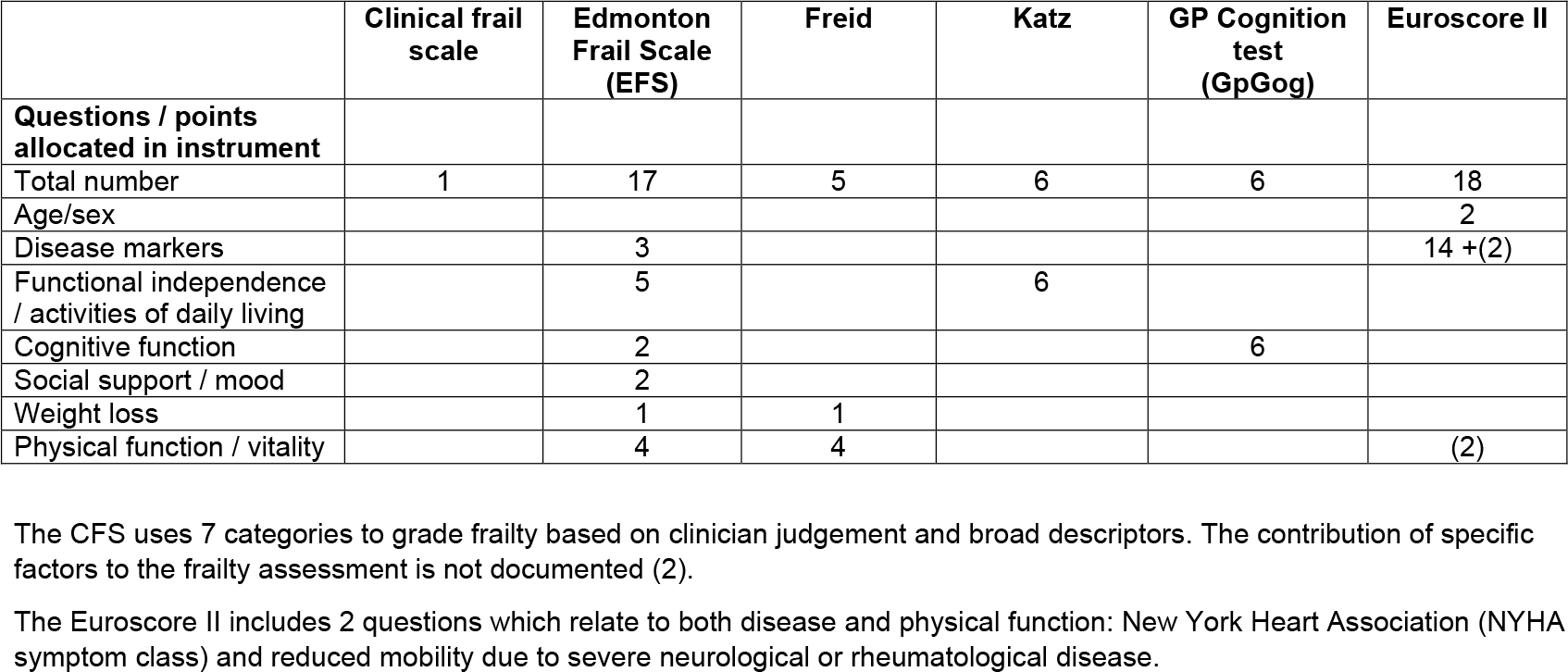
Comparison of instruments evaluated in this study.

|  | **Clinical frail scale** | **Edmonton Frail Scale (EFS)** | **Freid** | **Katz** | **GP Cognition test (GpGog)** | **Euroscore II** |
| --- | --- | --- | --- | --- | --- | --- |
| **Questions / points allocated in instrument** |  |  |  |  |  |  |
| Total number | 1 | 17 | 5 | 6 | 6 | 18 |
| Age/sex |  |  |  |  |  | 2 |
| Disease markers |  | 3 |  |  |  | 14 +(2) |
| Functional independence / activities of daily living |  | 5 |  | 6 |  |  |
| Cognitive function |  | 2 |  |  | 6 |  |
| Social support / mood |  | 2 |  |  |  |  |
| Weight loss |  | 1 | 1 |  |  |  |
| Physical function / vitality |  | 4 | 4 |  |  | (2) |

The CFS uses 7 categories to grade frailty based on clinician judgement and broad descriptors. The contribution of specific factors to the frailty assessment is not documented (2).
The Euroscore II includes 2 questions which relate to both disease and physical function: New York Heart Association (NYHA symptom class) and reduced mobility due to severe neurological or rheumatological disease.

After informed consent frailty assessments were undertaken >24 hours after admission when the patient was clinically stable. The CFS was completed by the clinical team caring for the patient. Random allocation was used to provide the clinical team with the results of the other frailty assessments either before or after completion of the CFS. Other frailty instruments were completed by a trained research nurse. All frailty assessments were then included in the medical record and were available to inform clinical care.

The CFS is based on data from the Canadian Study of Health and Aging and consists of 7 categories ranging from very fit to severely frail, with each category defined by a short description (10). The healthcare professional familiar with the patient scores the patient from an objective point of view. Because the CFS most closely reflects overall clinical judgment of frailty this scale was chosen to describe clinical risk factors and scores for other instruments by increasing frailty.

The Fried was derived with data from the Cardiovascular Health Study and defined by five criteria: unintentional weight loss in the past year, self-reported exhaustion, weakness measured by grip strength, slow walking speed, and low physical activity(11).

The EFS was developed for non-trained staff to assess frailty and includes 10 areas with a maximum score of 17 as the frailest individuals. The areas tested are mood, functional independence, social support, use of medication, nutrition, health perception, quality of life, continence, and two functional tests: the Clock test and the Timed get Up and Go test(12).

The Katz summarizes independent skills including bathing, dressing, going to the toilet, transferring, continence, and feeding. This measurement was developed as a predictor of need for nursing homes, alternative living, hospitalization, or home care(13).

The GPCog was designed to help GP’s detect dementia and to be quick to administer. The test includes the Clock test, short-term memory tests, and questions on managing self(14).

The Euroscore II was developed as a tool to assess the 30-day mortality risk in patients undergoing cardiac surgery. It is a score based on the severity of the underlying cardiovascular disease, the type of the planned surgery, and other co-morbidities(18).

### Linkage to registry and administrative data

The All of New Zealand, Acute Coronary Syndrome – Quality Improvement (ANZACS-QI) registry was completed in all cardiac centers and catheterization laboratories in New Zealand since 2015(19). The ANZACS-QI is a web-based system which captures data for the management of patients admitted to hospital with an acute coronary syndrome designed to provide information on the quality of care. Demographic and clinical information including age, sex, ethnicity, time at the index hospital admission, acute coronary syndrome diagnosis, current smoking, history of diabetes, hypertension requiring pharmacotherapy, prior cardiovascular disease, history of heart failure, Grace risk score, and percutaneous coronary intervention or referral for coronary artery bypass surgery during the index admission are recorded in ANZACS-QI. Frailty assessments were recorded in a research module linked to the ANZACS-QI registry.

All individuals in New Zealand in contact with the health system are assigned a unique number, the National Health Index (NHI). This can be linked to electronic health databases including the National Minimum dataset that records all hospital admissions by International Classification of Diseases (ICD) 10 codes and the Mortality Collection that records all deaths in New Zealand. The pre-specfied primary endpoints of the current study were all-cause mortality from the National Mortality dataset and hospitalization lasting > 10 days for any reason during the next year from the National Minimum dataset.

### Statistical analysis

Baseline clinical characteristics and summary statistics for the different frailty instruments are presented according to the 7 categories of increasing frailty on the CFS. Values were reported as means and medians with interquartile range. For other analyses responses on each of the instruments were grouped, when possible, in approximate quintiles of the study population, but also determined by the range of possible scores. The associations between the different frailty assessment tools and risk of death, and hospitalisation >10 days in the next year were analyzed using Cox proportional regression models, and reported by hazard ratios with 95% confidence intervals, adjusted by age and sex. Associations for CFS with mortality and hospitalisation were similar whether completed before or after access to other frailty scores, and were therefore reported together. Logistic regression was used because time to event was considered less relevant.

The performance of the different tools were evaluated by Harrell’s C statistics and the integrative discriminative index (IDI). IDI is the improvement in the difference in average predicted risks between the individuals with and without the outcome in the updated model, thereby the average improvement in sensitivity across all cutoffs (20). All statistical analyses were done by STATA version 15.1.

## Results

Baseline characteristics of the study population are displayed in Table 2 across 7 categories of the CFS. The percentage of women ranged from 22% in the severely frail group to 53% in the mildly frail group. The mean age was 76 years (interquartile range: 72-80), and was similar for men and women. In the total cohort 85 % were of New Zealand or other European descent, 6% New Zealand Maori, 6% Pacific Islanders, and 3% of Asian descent.

**Table 2.** Clinical characteristics of study population assessed during index hospitalization.

|  | All | Clinical Frail Scale |  |  |  |  |  |  |
| --- | --- | --- | --- | --- | --- | --- | --- | --- |
|  |  | Very fit | Well | Well comorbid | Vulnerable | Mildly frail | Moderately frail | Severely frail |
| **Number (%)** | 1174 (100) | 113 (10) | 182 (16) | 333 (28) | 280 (24) | 158 (13) | 99 (8) | 9 (1) |
| Women, % | 37 | 23 | 39 | 34 | 34 | 53 | 47 | 22 |
| Age | 76(72-80) | 76(73-80) | 75(71-79) | 76(72-79) | 76(73-80) | 78(73-83) | 77(74-83) | 75(74-83) |
| Women's age | 77(73-85) | 77(73-85) | 76(72-83) | 76(72-79) | 75(72-80) | 79(74-83) | 78(75-84) | 85(75-94) |
| **Ethnicity, N(%)** |  |  |  |  |  |  |  |  |
| NZ European | 810(70) | 80(72) | 120(66) | 239(72) | 187(67) | 110(71) | 68(72) | 6(67) |
| Other European | 174(15) | 23(21) | 31(17) | 38(12) | 49(18) | 20(13) | 11(12) | 2(22) |
| NZ Maori | 73(6) | 6(5) | 7(4) | 27(8) | 15(5) | 10(6) | 7(7) | 1(11) |
| Pacific Islands | 65(6) | 0(0) | 14(8) | 14(4) | 23(8) | 10(6) | 4(4) | 0(0) |
| Asian | 38(3) | 2(2) | 9(5) | 12(4) | 5(2) | 6(4) | 4(4) | 0(0) |
| **CVD + risk factors** |  |  |  |  |  |  |  |  |
| Current smoker | 74(7.3) | 2(2.1) | 7(4.3) | 28(9.8) | 23(9.5) | 8(6.1) | 6(7.2) | 0(0) |
| Diabetes | 292(29) | 20(21) | 27(17) | 80(28) | 84(35) | 42(32) | 35(42) | 4(50) |
| BMI >=30 | 254(31) | 13(17) | 42(33) | 77(32) | 73(38) | 25(24) | 22(34) | 2(29) |
| BMI<=20 | 25(3.1) | 2(2.6) | 4(3.2) | 2(0.8) | 5(2.6) | 7(6.7) | 5(7.7) | 0(0) |
| Prior CVD | 452(39) | 36(32) | 56(31) | 133(40) | 129(46) | 56(35) | 38(38) | 4(44) |
| Prior MI | 285(24) | 24(21) | 30(16) | 79(24) | 86(31) | 35(22) | 29(29) | 2(22) |
| Heart failure | 62(5) | 2(1.8) | 3(1.7) | 18(5.4) | 24(8.6) | 7(4.4) | 8(8.1) | 0(0) |
| COPD | 121(10) | 4(3.5) | 12(6.6) | 30(9.0) | 31(11) | 22(14) | 19(19) | 3(33) |
| Hemoglobin | 135(124-147) | 142(131-153) | 138(128-149) | 136(126-149) | 135(124-144) | 130(120-139) | 126(113-138) | 125(107-138) |
| Creatinine | 94(80-132) | 94(79-105) | 90(77-104) | 93(80-111) | 97(81-118) | 97(78-129) | 103(84-128) | 108(93-132) |

| Grace score | 3(1-6) | 3(1-4.5) | 2(1-4) | 3(1-5) | 3(1-7) | 3(1.5-6) | 4(2-7) | 17(3-24) |
| --- | --- | --- | --- | --- | --- | --- | --- | --- |
| Diagnosis |  |  |  |  |  |  |  |  |
| STEMI | 233(23) | 24(25) | 35(22) | 66(23) | 55(23) | 29(22) | 20(24) | 4(50) |
| Non-STEMI/UA | 716(71) | 68(71) | 111(68) | 214(75) | 169(70) | 91(69) | 59(71) | 4(50) |
| Not- ACS | 61(6) | 4(4) | 17(10) | 7(2) | 17(7) | 12(9) | 4(5) | 0(0) |
| Coronary revascularisation |  |  |  |  |  |  |  |  |
| PCI | 467(40) | 50(44) | 68(37) | 142(43) | 111(40) | 55(35) | 39(39) | 2(22) |
| CABG | 185(20) | 25(27) | 44(30) | 56(20) | 38(17) | 14(12) | 8(10) | 0(0) |

Numbers are median (interquartile range) or number (percent). N=number. NZ=New Zealand.
BMI = Body mass index in kg/m<sup>2</sup>, CVD = cardiovascular disease, COPD = chronic obstructive pulmonary disease, MI = myocardial infarction, ACS = Acute coronary syndrome, STEMI = ST elevation myocardial infarction, UA = unstable angina

According to the CFS 10% of study participants were very fit, 16% well, 28% well with comorbidities, 24% vulnerable, 13% mildly frail, 8% moderately frail, and 1% severely frail (Figure 1). The percentage of patients wirh diabetes, lower hemoglobin and higher creatinine levels increased with higher CFS. Patients with a high CFS score were less likely to undergo coronary revascularization.

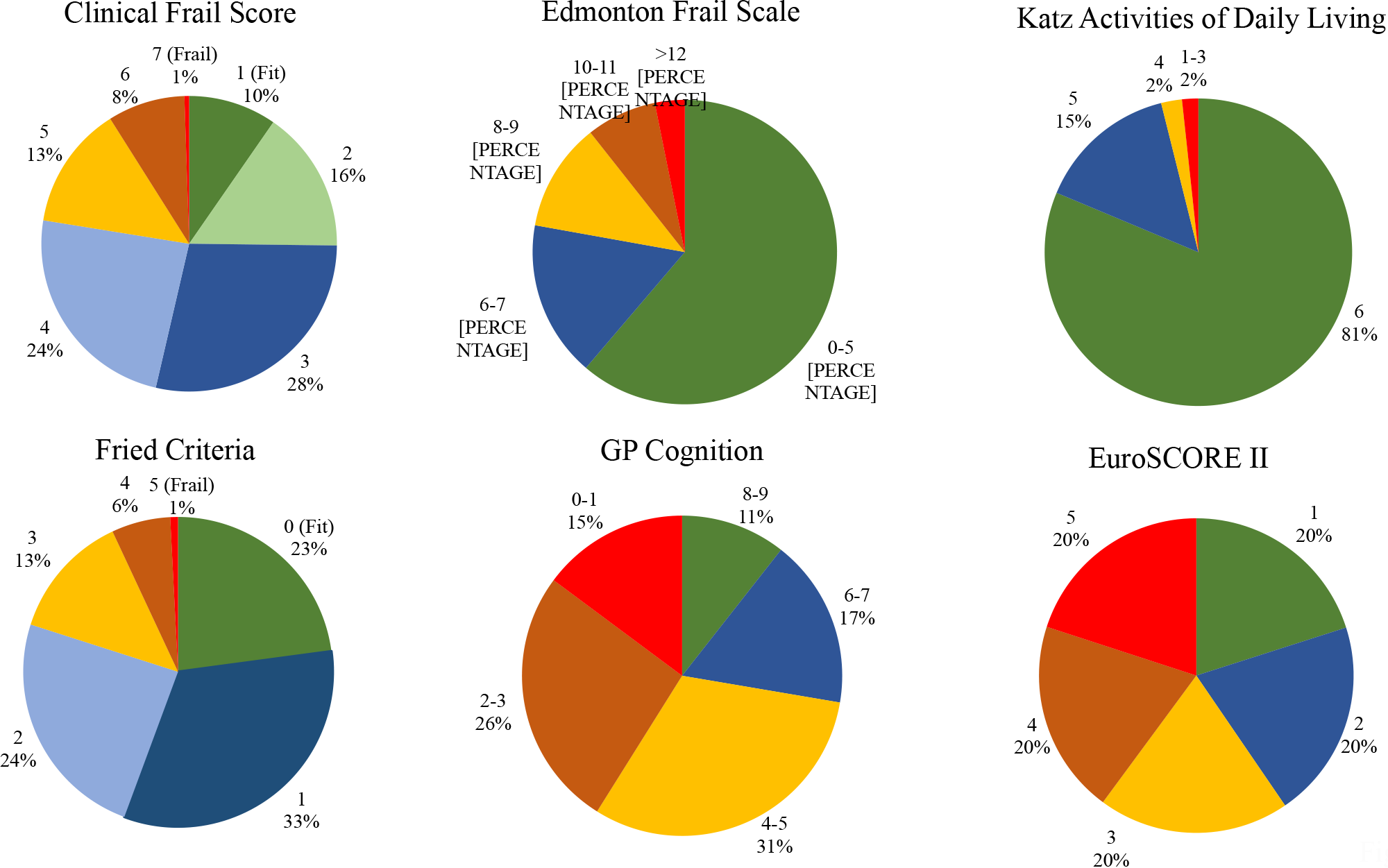

Summary measures for scores using the other instruments are displayed in table 3 by CFS score, and in figure 1. For the EFS 61% were classified as not frail, 17% vulnerable, 12% mildly frail, 7% moderately frail, and 3% severely frail. For the Katz 81% were independent in all 6 activities of daily living (ADL’s) and 15% in 5 ADLs. 23% of patients were frail according to the Fried (>=3 of 5 criteria present). The median score for the GPCog was 7 (interquartile range 5 to 8) and for the Euroscore II 2 (interquartile range 3 to 4).

**Table 3.** Scores for different frailty instruments and the EuroSCORE II by frailty based on the Clinical Frail Scale in patients admitted with a confirmed or suspected acute coronary syndrome.

|  |  | Clinical Frail Scale |  |  |  |  |  |  |  |
| --- | --- | --- | --- | --- | --- | --- | --- | --- | --- |
|  |  | All | Very fit | Well | Well comorbid | Vulnerable | Mildly frail | Moderately frail | Severely frail |
| Number (%) |  | 1174 (100) | 113 (10) | 182 (16) | 333 (28) | 280 (24) | 158 (13) | 99 (8) | 9 (1) |
| Frailty assessment |  |  |  |  |  |  |  |  |  |
| EFS | Median (IQR) | 4(2-7) | 2(1-4) | 3(2-5) | 4(2-6) | 5(3-7) | 7(4-9) | 9(6-11) | 12(10-13) |
|  | mean | 5.0 | 2.7 | 3.7 | 3.9 | 5.5 | 6.8 | 8.3 | 10.9 |
| Katz ADL | Median (IQR) | 6(6-6) | 6(6-6) | 6(6-6) | 6(6-6) | 6(6-6) | 6(5-6) | 6(5-6) | 4(3-6) |
|  | mean | 5.8 | 5.9 | 5.9 | 5.9 | 5.8 | 5.6 | 5.3 | 4.2 |
| Fried | Median (IQR) | 1(1-2) | 1(0-2) | 1(0-2) | 1(0-2) | 2(1-2) | 3(1-3) | 3(2-4) | 4(4-5) |
|  | mean | 1.5 | 1.0 | 1.0 | 1.1 | 1.7 | 2.4 | 2.6 | 4.5 |
| GP Cognition | Median (IQR) | 7(5-8) | 8(6-8) | 7(5-8) | 7(6-8) | 7(5-8) | 6(5-8) | 6(4-8) | 5(1-7) |
|  | mean | 6.5 | 7.3 | 6.7 | 6.8 | 6.4 | 6.0 | 5.5 | 4.3 |
| EuroSCORE II | Median (IQR) | 2(2-4) | 2(1-3) | 2(1-3) | 2(1-4) | 3(2-5) | 3(2-6) | 4(2-6) | 10(4-13) |
|  | mean | 3.6 | 2.3 | 2.8 | 3.0 | 3.8 | 5.0 | 5.2 | 10.5 |

Numbers are mean (interquartile range) or number (percent). N=number. EFS=Edmonton Frail Scale. ADL=Activities of Daily Living. GP=General Practitioner. EuroSCORE= The European System for Cardiac Operative Risk Evaluation.

### Risk of mortality according to frailty scores

During a median follow-up of 5.1 (IQR: 4.6-5.5) years there were 353 deaths from all causes (29%) There were graded increases in mortality risk with increasing ‘frailty’ assessed on all the instruments (Figure 2). For the EFS there was a stepwise increase in the hazard ratio (HR) for allcause mortality with increasing frailty, which was 5.0 (95% CI: 3.4-7.4) for patients with higher scores from 9-17 (n=197) compared to low scores (score 0-2, n=331).

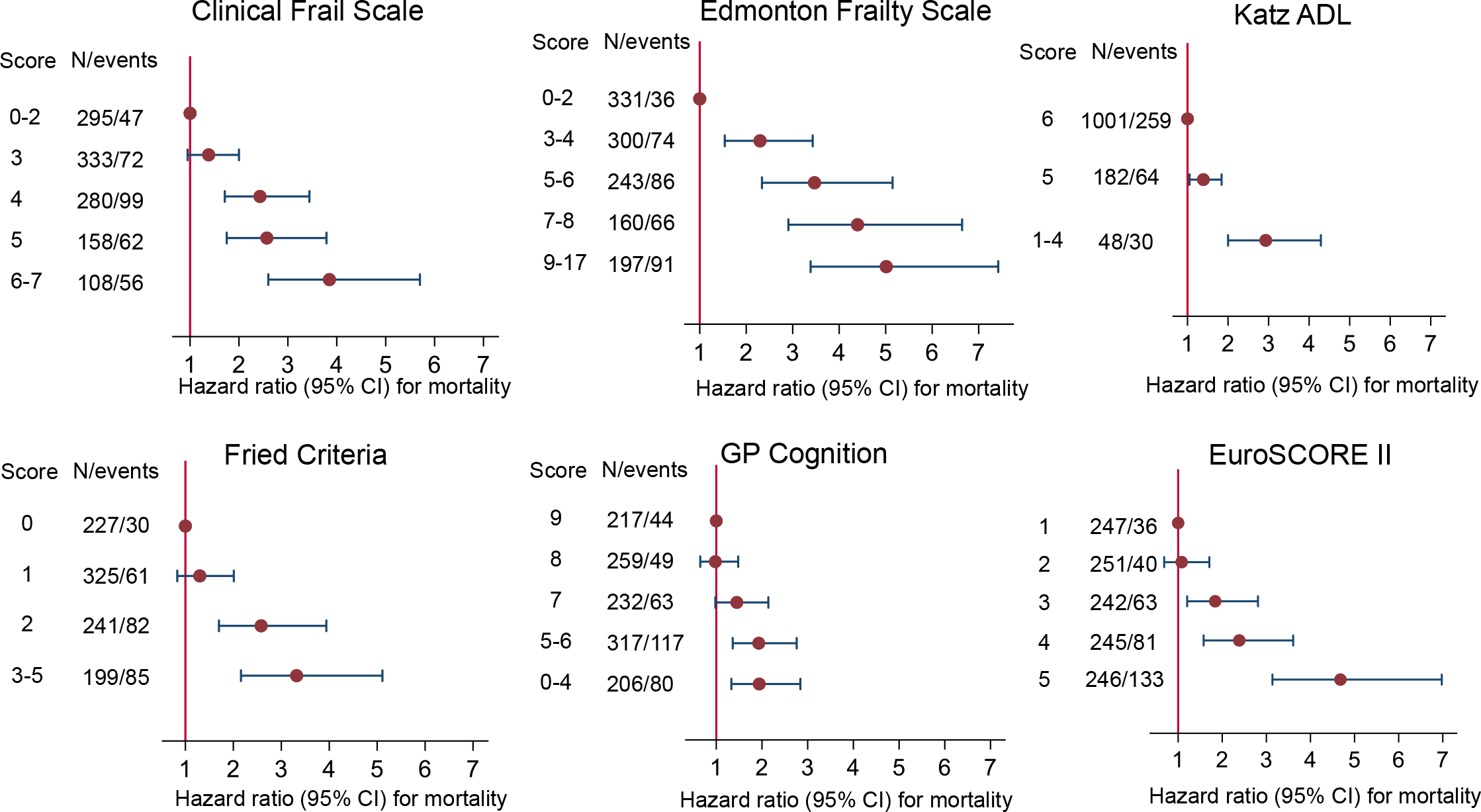

During 12 months after the index hospitalization 267 patients (22%) were hospitalized for > 10 days. HRs for hospitalization >10 days in the next year for each of the frailty tools are presented in Figure 3. There were graded increases in the HR for hospitalization > 10 days in the next year for each of the tools, which were strongest for the EFS and Fried.

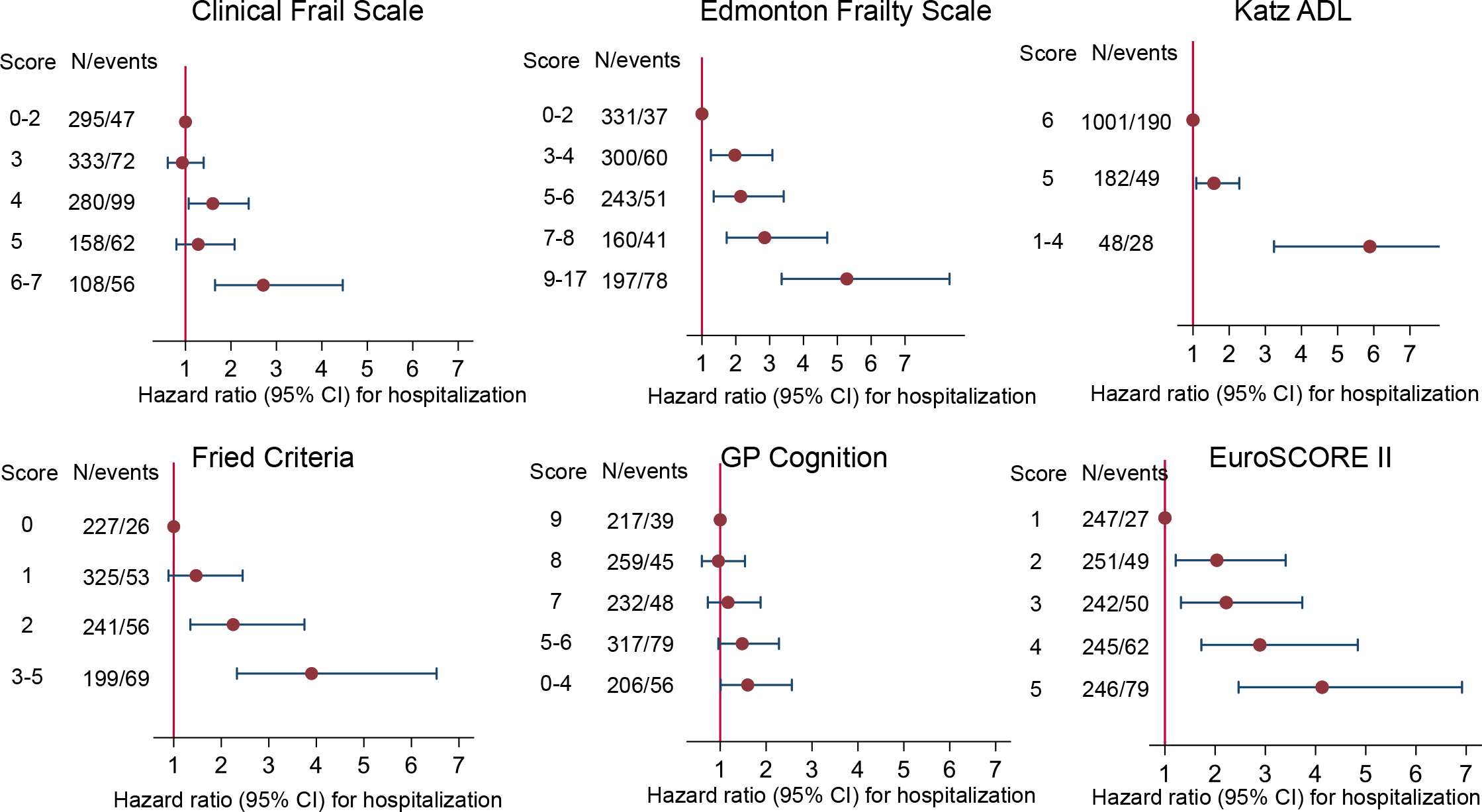

The C-statistics for both mortality and hospitalization are compared for the different tools in Table 4. The discrimination of all-cause mortality according to Harrell’s C-index was 0.663 (95%CI: 0.635-0.692) for the EFS, 0.648 (0.614-0.683) for the Fried, 0.641 (0.610-0.671) for the CFS, 0.608 (0.578-0.638) for the GPCog, and 0.593 (0.560-0.626) for Katz (Table 2). The C-statistics for hospitalization >10 days in the next year were higher for EFS 0.649 (0.611-0.687) and the Fried 0.628 (0.585-0.672) compared to other instruments. The EFS was the tool with the highest Cstatistic for both mortality and hospitalisation.

**Table 4.** Frailty assessment score’s discrimination for mortality and hospitalization in patients with acute coronary syndrome.

| Frailty assessment | Mortality |  |  | Hospitalization |  |
| --- | --- | --- | --- | --- | --- |
|  | C-statistic | 95% CI |  | C-statistic | 95%CI |
| Clinical Frail Scale | 0.641 | 0.610-0.671 |  | 0.584 | 0.544-0.623 |
| Edmonton Frailty Scale | 0.663 | 0.635-0.692 |  | 0.649 | 0.611-0.687 |
| Fried Criteria | 0.648 | 0.614-0.683 |  | 0.628 | 0.585-0.672 |
| General Practitioner Cognition Test | 0.608 | 0.578-0.638 |  | 0.552 | 0.513-0.590 |
| Katz Independence in ADL | 0.593 | 0.560-0.626 |  | 0.602 | 0.560-0.644 |
| EuroSCORE II | 0.654 | 0.622-0.686 |  | 0.589 | 0.550-0.629 |

Analyses were adjusted by age and sex. CI=confidence interval. ADL=activities of daily living. EuroSCORE= The European System for Cardiac Operative Risk Evaluation.

The improvement in risk discrimination when adding additional information to another tool, measured using the integrated discrimination improvement (IDI), are compared in Table 4. Of the frailty tools the EFS added the most discrimination for both frailty and hospitalization. When the EFS was performed first, other frailty tools added less predictive information. The Euroscore II added more predictive information for mortality when added after all frailty tools. The EFS and Euroscore II added a similar amount of prognostic information both independently and when used together (Table 5).

**Table 5.** Integrated Discrimination Improvement (IDI) for A) all-cause mortality and B) hospitalization by adding information from different frailty scales to initial assessment tool

| Initial assessment |  |  |  |  |  |  |
| --- | --- | --- | --- | --- | --- | --- |
|  | +Clinical FS | +Edmonton FS | +Fried Criteria | +Katz ADL | +GP Cog | +EuroSCORE II |
| Clinical Frail Scale | - | 0.0387<br>(p<0.0001) | 0.0291<br>(p<0.0001) | 0.0099<br>(p=0.003) | 0.0146<br>(p=0.0001) | 0.0602<br>(p<0.0001) |
| Edmonton Frailty Scale | 0.0139<br>(p=0.0001) | - | 0.0106<br>(p=0.002) | 0.0021<br>(p=0.14) | 0.0073<br>(p=0.006) | 0.0481<br>(p<0.0001) |
| Fried Criteria | 0.0208<br>(p<0.0001) | 0.0245<br>(p<0.0001) | - | 0.0009<br>(p=0.4) | 0.0098<br>(p=0.005) | 0.0599<br>(p<0.0001) |
| Katz ADL | 0.0441<br>(p<0.0001) | 0.0593<br>(p<0.0001) | 0.0569<br>(p<0.0001) | - | 0.0224<br>(p<0.0001) | 0.0776<br>(p<0.0001) |
| GP Cognition Test | 0.0479<br>(p<0.0001) | 0.0630<br>(p<0.0001) | 0.0527<br>(p<0.0001) | 0.0210<br>(p<0.0001) | - | 0.0832<br>(p<0.0001) |
| EuroSCORE II | 0.0259<br>(p<0.0001) | 0.0425<br>(p<0.0001) | 0.0280<br>(p<0.0001) | 0.0149<br>(p=0.0005) | 0.0219<br>(p<0.0001) | - |

B.
| Initial assessment |  |  |  |  |  |  |
| --- | --- | --- | --- | --- | --- | --- |
|  | +Clinical FS | +Edmonton FS | +Fried Criteria | +Katz ADL | +GP Cog | +EuroSCORE II |
| Clinical Frail Scale | - | 0.0350<br>(p<0.0001) | 0.0282<br>(p<0.0001) | 0.0185<br>(p=0.0002) | 0.0024<br>(p=0.1) | 0.0136<br>(p=0.0007) |
| Edmonton Frailty Scale | 0.0005<br>(p=0.5) | - | 0.0088<br>(p=0.01) | 0.0080<br>(p=0.01) | 0.0005<br>(p=0.4) | 0.0078<br>(p=0.007) |
| Fried Criteria | 0.0021<br>(p=0.1) | 0.0116<br>(p=0.004) | - | 0.0080<br>(p=0.03) | 0.0007<br>(p=0.3) | 0.0137<br>(p=0.003) |
| Katz ADL | 0.0073<br>(p=0.005) | 0.0300<br>(p<0.0001) | 0.0315<br>(p<0.0001) | - | 0.0045<br>(p=0.03) | 0.0145<br>(p=0.0004) |
| GP Cognition Test | 0.0142<br>(p<0.0001) | 0.0463<br>(p<0.0001) | 0.0391<br>(p<0.0001) | 0.0283<br>(p<0.0001) | - | 0.0190<br>(p<0.0001) |
| EuroSCORE II | 0.0090<br>(p=0.003) | 0.0409<br>(p<0.0001) | 0.0277<br>(p<0.0001) | 0.0256<br>(p<0.0001) | 0.0063<br>(p=0.01) | - |

## Discussion

### Associations between frailty scores and mortality and hospitalisation

Increasing frailty was associated with higher all cause mortality and prolonged hospitalisation for all the instruments assessed. This was most clear for the EFS, which also identified differences in mortality risk between patients who did not meet criteria for frailty according to the Fried or Katz scores. These observations indicate that in a population of ACS patients predominantly aged >70 years a simple categorisation of frailty as present or absent provides less prognostic and clinical information compared to assessment of frailty on a graded scale.

Three frailty instruments, the EFS, Fried, and CFS had similar C-statistics for mortality compared to the Euroscore II, and added incremental prognostic information when combined with the Euroscore II. The Euroscore II uses established disease based risk factors to estimate 30-day mortality risk following cardiac surgery, and also predicts longer term mortality (21). Because the frailty tools and the Euroscore II provide different information, the incremental prognostic value when Euroscore II was used with a fraily tool such as the EFS was greater than using 2 different frailty tools.

The CFS, which is based on clinician judgment was the simplest frailty assessment in this study. For mortality the predictive performance of the CFS was slightly less than the EFS, and similar to the Fried. This observation supports the ability of clinical judgment to estimate prognosis related to frailty as a simpler alternative to more structured frailty instruments. However, the CFS was less predictive of hospitalization than the EFS or Fried, and does not include a formal assessment of different dimensions of frailty such as low physical activity, weakness and weight loss, which may be useful to guide clinical care.

The Katz was less strongly predictive of mortality than other frailty instruments. In this study 81% of the participants had no, and 96% ≤ one limitation of daily living assessed by Katz. The Katz is therefore not likely to be a useful as an initial frailty assessment tool for the majority of older acute coronary syndrome patients. The GPCog identified a broad range of scores, but was a weaker predictor of both mortality and hospitalization compared to the CFS, EFS, and Fried. The GPCog assesses cognitive function, which may not predict mortality as strongly as other dimensions of ‘frailty’ such as weight loss, weakness and low physical activity.

### Comparison with other studies

Previous studies have reported that frail patients with cardiovascular disease are more likely to suffer adverse outcomes (15). In a meta-analysis which evaluated associations between frailty and all-cause mortality after myocardial infarction (22), the combined hazard ratio for frail versus non-frail patients by the Fried was 2.8 (95%CI: 1.1-2.5)(23,24). The CFS has been associated with in-hospital mortality, short and long-term all-cause mortality, longer hospitalization, and risk of future hospitalization (25-30). The EFS has been associated with all-cause mortality in patients with an acute coronary syndrome (31-33). Frailty according to the Katz was associated with a higher risk of postoperative complications, in-hospital mortality, institutional discharge, and reduced survival in 3826 patients undergoing cardiac surgery (34). Reduced cognitive function identified using the GPCog has been associated with all-cause and cardiovascular mortality in mid- and late-life (35-38).

However, few previous studies have directly compared different frailty instruments in a large cohort of patients with cardiac disease. One study reported increased mortality and hospitalization for frail compared to non-frail patients assessed using the Fried, the EFS, and the CFS in 174 acute coronary syndrome patients, but with limited analysis of differences between instruments(16). Previous studies did not directly compare discrimination of different frailty instruments for adverse clinical outcomes in a large cohort of acute coronary syndrome patients.

### Clinical implications

The clinical value of different frailty assessment tools depends on the information obtained from individual questions, and the association between the overall ‘frailty score’ and adverse clinical events. The CFS provides less information on reasons for frailty because it uses a simple scale with scoring based on clinical judgment alone. Other frailty instruments score responses to specific questions –but there are large differences between frailty tools (Table l). The Fried assesses the frailty phenotype which includes wasting, weakness, and low physical activity. The GPCog is a formal assessment of cognitive function, which in itself is important for clinical decisions, but does not assess frailty. Strengths of the EFS are that it evaluates several dimensions of ageing including physical and cognitive function, weight loss, and activities of daily living, each of which may be relevant to clinical care.

A frailty assessment may provide information relevant to clinical decisions in patients with a broad range of cardiovascular diseases. For some clinical decisions, including an invasive stategy in acute coronary syndrome patients, aortic valve replacement, implanted defibrillators, cardio-thoracic surgery, and costly medical treatments, a frailty assessment may better inform the balance of benefits and risks of the proposed intervention (5,15,34). In the current study frail patients were less likely to be referred for PCI or CABG. Evaluating frailty on a graded scale is an advantage if the clinically relevant level of frailty which influences decisions varies by treatment or indication. A frailty assessment can also guide other aspects of clinical care, such as targeting assistance for specific difficulties with daily living, encouraging exercise in patients with weakness and low functional capacity, and modifying treatment plans for patients with cognitive impairment. Measuring frailty on a continuum between good health and advanced frailty is also relevant to prevention, which includes maintaining or increasing physical activity, healthy eating and good medical care.

### Study limitations

Frailty assessments were undertaken by experienced cardiac nurses at each participating hospital after brief training. The use of these instruments was therefore similar to usual clinical care.

Variation in evaluations between assessors was not determined. Within the same study population the discriminatory value of different frailty instruments can be directly compared. However it is possible results vary for different countries and ethnicities, and for patients with different medical problems. Strengths of the study include the large number of patients with standard information collected for the different frailty assessment tools, and complete follow-up for outcomes from comprehensive national administrative data.

## Data Availability

The Data will be made available on request

## Conclusion

In acute coronary syndrome patients mostly aged >70 years increasing frailty scores was associated with all cause mortality for all the tools evaluated. The Edmonton Frailty Scale provides a graded assessment of the level of frailty, information on factors contributing to frailty, and predicted allcause mortality and hospitalization as well as or better than other frailty instruments.

